# Maintenance of Heterochromatin links Chromatin Modifiers and Neurodevelopment in Autism Spectrum Disorder

**DOI:** 10.1101/2023.10.10.23296804

**Authors:** Michael R. Garvin, David Kainer

## Abstract

Autism spectrum disorder (ASD) is a highly heritable and highly heterogeneous neuropsychiatric condition whose cause is still unknown because there are no recurrent genes found among diagnosed individuals. One of the most common functional properties of the many reported risk-genes for autism is “chromatin modification” but it is not known how this biological process relates to neurodevelopment and autism. We recently reported frequent, recurrent genomic structural variants (SVs) in two cohorts of individuals with autism that were identified using non-Mendelian inheritance (NMI) patterns in family trios. The genes harboring the SVs participate in neurodevelopment, glutamate signaling, and chromatin modification, confirming previous reports and providing greater detail for these processes in ASD. The majority of these ASD-associated SVs (ASD-SV) were found in non-coding regions of the genome and were enriched for expression quantitative trait loci (eQTL) suggesting that gene dysregulation results from these genomic disruptions rather than alteration of proteins. Here, we intersect the ASD-SV from our earlier work with different gene regulatory and epigenetic multiomic layers to understand how they may function to produce autism. Our results indicate that the core of ASD resides in the dysregulation of a process called RNA-induced Initiation of Transcriptional gene Silencing (RITS) that is meant to maintain heterochromatin and produces SVs in the genes within these chromosomal regions, resulting in alterations in brain development. This finally links reported ASD-risk genes involved in chromatin remodeling with neurodevelopment. In addition, it may explain the role of *de novo* mutations in ASD and provide a framework for more accurate diagnostics and endophenotypes.

## Introduction

There is now considerable effort within the medical community to develop and adopt a Precision Medicine approach whereby a patient’s genetic, environmental, and lifestyle information is used to tailor treatment at the level of the individual ^1^. The genetic component of this effort has the potential to be highly impactful if genomic mutations that cause biomedical traits of interest can be identified, because those changes represent crucial diagnostic and treatment possibilities.

As we and others have reported, autism spectrum disorder (ASD) is highly heritable (50-80% is attributed to inherited genetic variants), yet there is no recurrent gene or mutation that provides insight into the functional causes of autism or explains the heterogeneity ^2–5^. We recently revealed that considerable genetic and biological underpinnings of ASD reside in genes that harbor genomic structural variants (SV) of large effect that are difficult to identify with widely used sequencing technology but can be indirectly identified using non-Mendelian inheritance (NMI) patterns in family trios ^5^. By focusing on the subset of these SVs that were found in much higher frequency in people with ASD we identified the gene *ACMSD* as statistically significantly associated with non-verbal forms of autism. This provided important biological insights into ASD because the ACMSD enzyme functions to convert a toxic intermediate in the tryptophan salvage pathway (quinolinic acid) to a neuroprotective compound (picolinic acid). This pathway has been linked to several neuropsychiatric conditions including ASD, but that specific enzyme had not. Similarly, using our method to identify SVs, we predicted and then confirmed abnormal splicing of the glutamate receptor subunit *GRIK2* in a subset of individuals with autism. This gene had also been linked to neuropsychiatric disorders including obsessive compulsive disorder and autism in those of Korean ancestry ^6,7^, and our work provided the molecular mechanism and specific genetic location as it relates to ASD. Using our entire matrix of SVs, we were able to identify subtypes of autism by clustering ASD cases according to their genomic SV profiles.

Toward the goal of further disentangling the complex heterogeneity of ASD, in this study we take a multiomic approach to understand the functional genomics of the entire set of SVs we reported previously. Our results indicate that the most important and frequent ASD-related SVs are located in heterochromatin-rich regions of the genome, and they are enriched in binding sites for transcription factors that regulate heterochromatin itself. This corresponds well with the fact that regulation of chromatin is one of the most commonly reported functions of genes known to be mutated in individuals with autism ^8–13^. For example, of the nearly 800 risk genes for ASD listed by the Simons Foundation Autism Research Initiative (SFARI), at least 50 participate in the regulation of chromatin. Despite the obvious importance of this function in autism, there is currently no clear understanding or hypothesis of how genes involved in chromatin modification link to autism beyond generally affecting the expression of genes or epigenetic modifications to the DNA ^14^.

By integrating the set of ASD-related SVs with regulatory genomic data such as eQTLs, heterochromatin, methylation, and transcription factor binding sites, for the first time we provide an explanation for the link between chromatin remodeling and brain development in autism. Our results indicate that the disorder initially derives from SVs that cause dysregulation of genes that comprise the RNA-induced Initiation of Transcriptional gene Silencing (RITS) and SWI/SNF (BAF) chromatin remodeling complexes. These two multi-subunit complexes are meant to precisely balance suppression and expression of genomic regions that are rich in repetitive elements and transposons, which can be mutagenic if not tightly regulated. ASD results from the imbalance of this system because it produces SVs in the developmental genes that also happen to reside in these regions. A dysfunctional RITS and chromatin modification system would also explain the observation of *de novo* mutations in many cases of ASD because loss of protective heterochromatin would result in a higher mutation rate.

## Materials and Methods

### Structural Variant dataset

For this study we used a set of SNP-array loci that tag putative structural variants in ASD cohorts from the MIAMI and AGPC GWAS studies ^5^. Briefly, we previously used these loci to detect SVs using patterns of non-mendelian inheritance (NMI) in the family trios within these cohorts. We showed that the NMI pattern often indicated erroneous genotyping due to genomic features, such as SVs, disrupting the binding of the array probe rather than due to random error. These NMI loci were stringently filtered to a subset, known as ASD-SVs, that were not already flagged as SVs in the broader population, and were found at much higher frequency in the ASD cohorts. The genes harbouring this set of ASD-SVs were shown to be heavily enriched for brain development and chromatin regulation functions, and could be used to identify genetically distinct clusters of ASD cases.

### Overlap of ASD-SVs with genomic features

Here, in order to understand the genomic context of the ASD-specific SV loci, we used only those that were found in 15% or more of individuals in both studies used in our previous report ^5^, leaving 2,468 array SNPs that identified potential genomic structural variation (Figure 1, Table S1). For genomic region enrichment analysis, we generated a *bed* file of intervals flanking +/- 1kb from each of the 2,468 NMI SNPs. The majority of the ASD-SVs we detected and reported previously ^5^ were in non-coding space, which suggested that they affect regulatory or epigenetic mechanisms rather than protein function. Therefore, we intersected these intervals with the eQTL, heterochromatin, RepeatMask, CpG, ORegAnno, and Chip-seq data from RepMap Atlas ^15^ available from the UCSC Table browser using the Galaxy platform ^16^ because they represent different aspects of these non-coding processes. For the eQTL section, rather than the 2kb overlap window, we queried for an exact match of the SNP locus and for the tissue-specific analysis, we binned tissues by organ (e.g. brain) where possible.

**Figure 1.**
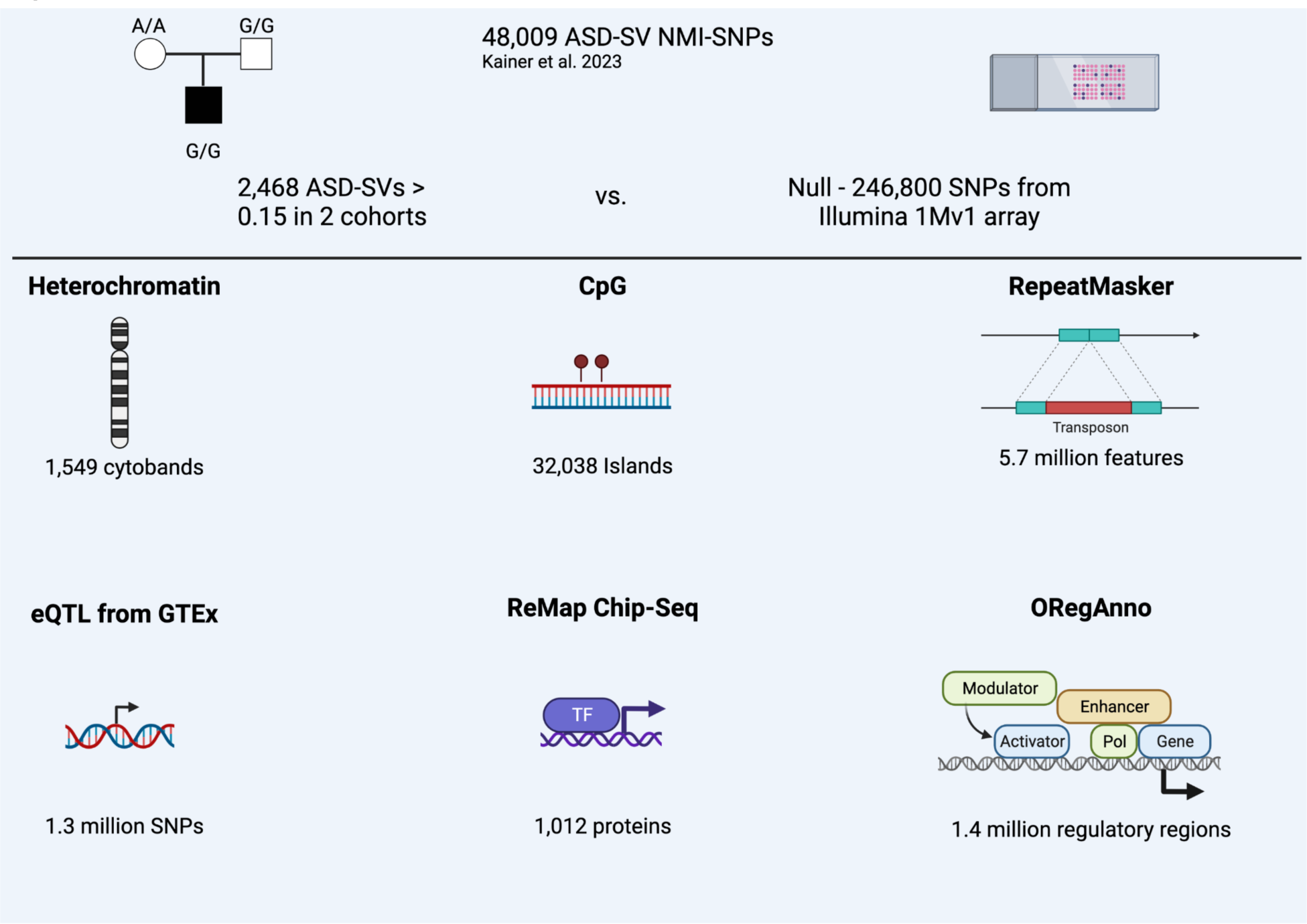
Overview of the analyses. ASD-SVs tagged with Non-Mendelian inherited (NMI) SNPs and found in greater than 15% of both the Miami and AGPC cohorts were extracted from the full database, which produced 2,468 SNPs for further analysis. We randomly sampled the same number 100 times from the remaining ∼ 900,000 SNPs on the Illumina array used for those studies and then intersected them with genomic features available on the UCSC table browser. Created with BioRender.

As a control, we did the same for 100 sets of 2,468 SNPs randomly sampled (without replacement) from the loci listed on the Illumina 1Mv1 array used for the studies that generated the original data analyzed in our report ^17,18^. Null estimates were generated from the 100 sets of 2,468 randomly sampled loci and compared to the ASD-specific set using a chi-square test and Bonferroni correction. Counts of any feature that were five or less at a site across a dataset were removed due to the inaccuracy of the chi-square test for these low values.

### Gene Ontology enrichment

For the Gene Ontology enrichment, we were interested in the genes that are regulated by the transcription factors from the Chip-seq ReMap project because they were significantly enriched for the ASD-SV. The genes regulated by these transcription factors may be causal of autism because the ASD-SV that overlap their regulatory region from ReMap are frequent (f > 0.15) in two ASD cohorts. In order to ensure we captured the biological significance of these genes, we expanded the list to include ASD-SVs that were greater than 5% in both ASD cohorts and overlapped transcription factor binding sites found to be significantly enriched in our Chip-seq ReMap analysis. Many of these ASD-SV-tagging SNPs (N=1041, Table S2a) are not in genic space and therefore we used several methods to assign them to a gene. First, we assigned to genes using the SNPnexus portal for Entrez ^19^, then with an exact rsID match in GTEx, and finally with a bed file intersect with ORegAnno 3.0 ^20^ (Table S2b). The data used for the analyses described in this manuscript were obtained from UCSC Table Browser and the GTEx Portal on 02/26/2023. The protein coding genes were submitted to Gene Ontology (geneontology.org/) for biological process (BP) term enrichment with a significance threshold FDR < 0.05.

## Results

### Heterochromatin

The heterochromatin feature on the UCSC Table browser identifies 7 regions: centromeric (acen) no heterochromatin (gneg), variable-length (gvar), and four levels of Giesma staining ranging from low to high (gpos25, gpos50, gpos75, gpos100). The SNPs that tagged ASD-SV were significantly enriched for the greatest staining intensity, i.e. the highest level of heterochromatin (gpos100, *p* < 1.4 x 10^-9^) and centromeric (acen, *p* < 8.3 x 10^-3^) compared to the null sets of SNPs. They were significantly lower in euchromatin (gneg, *p* < 1.4 x 10^-3^), Table S3.

### Repeat Mask

The repeat mask database lists 275 repetitive sequences taken from the RepBase update assembled by the Genetic Information Research Institute (www.girinst.org) that are binned into 8 classes ^21^. The ASD-SV tagging SNPs were significantly enriched in 5 classes (DNA transposons, LINE elements, satellite DNA, simple repeats, and snRNAs, *p* < 8.1 x 10^-4^). There were no differences between the ASD-SV tagging SNPs and the random SNPs for long terminal repeats, low complexity regions or SINE elements (Table S4). When looking within each class, the significant LINE elements are type L1 and not L2, and the simple repeats are for AT or TA repeats (Table S5). Type L1 LINE elements are younger than L2 and some are still active in the genome. They are responsible for retrotransposition of non-autonomous retrotransposons (e.g. *Alu*) as well as non-coding RNA and mRNA to produce pseudogenes ^22^.

### Gene Regulation

We tested for several regions associated with gene regulation and the ASD-SVs. CpG sites are often found in promoter regions and can result in gene regulation changes via differing methylation patterns as a result of mutations. There was no significant difference in the number of CpG sites between randomly chosen SNP loci and ASD-SV tagging SNPs (Table S6). The Open Regulatory Annotation database (ORegAnno) is an open-source database for gene regulation curation and is available via the UCSC Table Browser. For the ASD-SV SNPs, there were fewer than expected overlaps of annotated ORegAnno sites overall (all transcription factor binding sites) as well as coding genes regulated by the transcription factors compared to randomly chosen SNPs (*p* < 8.8 x 10^-21^ and *p* < 6.0 x 10^-4^, respectively), suggesting that altered gene regulation is not the functional outcome of these mutations (Table S6). However, in agreement with our previous report, they were enriched for eQTLs (*p* < 3.5 x 10^-18^). Here, we expanded this analysis to include tissue-specific tests and found that eQTLs were significantly enriched in the brain (*p* < 6.4 x 10^-6^) and lung (*p* < 1.2 x 10^-3^). Finally, the Chip-seq analysis with the 1,012 transcription factor binding sites in the ReMap Atlas indicate that the majority of these sites are either not different between the ASD-SV and control sets of SNPs, or are found significantly less frequently in the ASD-SV tagging SNPs. However, three transcription factor binding sites were enriched in the ASD-SV tagging SNPs after Bonferonni correction: SATB1 (*p* < 2.7 x 10^-25^), SRSF9 (*p* < 1.2 x 10^-8^), and NUP98-HOXA9 (*p* < 1.1 x 10^-6^), Table S8.

### Gene Ontology

To determine if dense heterochromatin harbored genes of related functions, we performed a biological enrichment test on the coding-genes found in the gpos100 regions identified in the UCSC Table Browser (N = 1194) and compared those to the same number of randomly sampled genes from euchromatin (nine sets after sampling without replacement, Table S9). The genes found in dense heterochromatin were significantly enriched for specific functions related to immune response (e.g. *natural killer cell activation involved in immune response,* 10.3-fold enriched) and brain development (e.g. *synaptic membrane adhesion*, 6.2-fold enriched) whereas seven of the nine euchromatin gene sets (N = 1194 genes each) were found to have no significant enrichments. Two of the nine were enriched for the broad and uninformative category of *cellular process*, 1.1-fold enriched.

We next wanted to know the function of the genes that are regulated by SABTB1, SRSF9 and NUP98-HOXA9 transcription factors but with binding sites that are potentially disrupted by ASD-SVs. There were 585 coding genes that were assigned to the ASD-SV tagged SNPs (Table S2b) within those binding sites. Sixty of the 585 were listed in GO terms associated with chromatin, leaving 525 for enrichment analysis that represent non-chromatin processes related to ASD. The GO submission generated 36 significantly enriched top-level terms for Biological Processes (FDR < 0.05, Table S10). As with our previously published result ^5^, dendritic spine development and glutamate receptor signaling featured prominently here, plus developmental processes involving the retinal and olfactory systems. The process *retinal ganglion cell axon guidance* was more than 10-fold enriched and *olfactory bulb development* more than 14-fold. Interestingly, the analysis identified 11 genes representing a 4.4-fold enrichment for female gonad development, which might potentially explain sex differences in diagnosed cases of ASD (Table S10).

## Discussion

Here we used a set of ASD-related SVs we had identified in our previous report ^5^ and analyzed them in a genomic context, with the results indicating that the principal dysregulated process in ASD is the maintenance of heterochromatin. We show significant over-representation of the ASD-SVs in heterochromatin and under-representation in euchromatin. The ASD-SVs are also more often found overlapping features known to be associated with heterochromatin such as ALR-alpha satellite DNA, transposons, small nuclear RNAs, and simple repeats. Although genes involved in chromatin remodeling have been reported in autism previously and are present in our ASD-SV (Table S11) ^23–25^, our results here indicate that the key process is specifically chromatin remodeling as it relates to the *regulation of heterochromatin* rather than gene regulation or epigenetics in the broader sense. Below we provide a clearer and more holistic understanding of how this biological process relates to downstream neurodevelopment.

### Heterochromatin and RNA-induced initiation of transcriptional gene silencing (RITS)

Heterochromatin is generated by epigenetic modifications of the histone proteins around which the DNA is wrapped, resulting in compaction and inaccessibility of the genome to transcription or replication. This is necessary because they harbor repetitive sequences or transposable elements that could generate SVs and genomic disruptions through either active transposition or non-homologous recombination ^26,27^. However, these regions are not simply “junk” but also harbor genes that code for protein and must be accessible to transcriptional machinery at certain times. This likely drove an evolutionary response to repress parasitic genomic elements such as transposons, but simultaneously allow for the tightly controlled expression of genes that are important for development and cellular differentiation in those same regions ^28,29^.

Amazingly, even though only 1% of the genome encodes for proteins, nearly three-quarters of the genome is transcribed into RNA ^30,31^. Much of this material is used for the regulation of heterochromatin, which is often characterized as facultative heterochromatin and constitutive heterochromatin. Facultative heterochromatin is typically associated with methylation of histones at the lysine 27 residue (H3K27me3) whereas constitutive heterochromatin is associated with tri-methylation of histones at lysine residue 9 (H3K9me3) and is fully methylated throughout the cell cycle, although these terms are fairly plastic ^32^. H3K27 is regulated by the polycomb repressive complex (PRC) whereas H3K9me3 is maintained by a protein complex called RNA-induced initiation of transcriptional gene silencing (RITS) that results in compaction of DNA and inaccessibility by transcription machinery. The system was originally elucidated in yeast (*S. pombe*) and later in mammals, and therefore the gene nomenclature can be confusing (Table S11).

Access to these regions begins with the removal of methylation at H3K9 residues by demethylases (i.e. erasers) that open the local genome to transcription. Reversion to a quiescent state is accomplished by converting the transcribed genomic regions to double stranded RNA oligos by DICER, which are then used to target the RITS complex back to the initial location of transcription so that H3K9 methylases (i.e. writers) can re-methylate the histones (Figure 2). In the context of disease, it is important to note that if this process is not tightly controlled, heterochromatin spreads into neighboring genomic regions, causing their suppression. This is called position effect variegation (PEV), and was discovered as the causative process of the white-eye phenotype in *Drosophila* because a transposition in that mutant resulted in heterochromatin being placed next to the *white* gene, causing it to be silenced ^33^.

**Figure 2.**
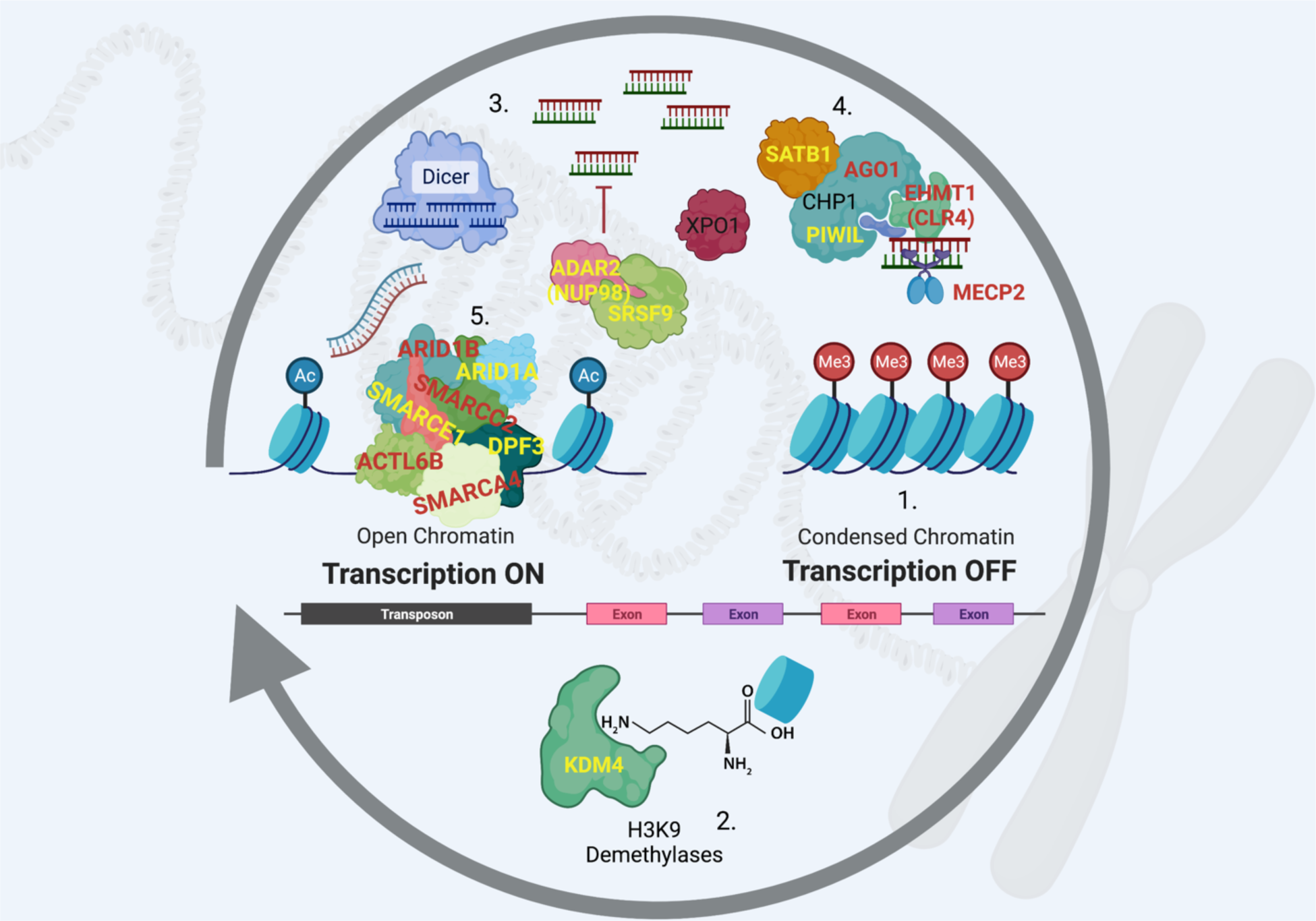
RNA-induced initiation of transcriptional gene silencing (RITS) and chromatin remodeling. 1. Constitutive heterochromatin is maintained in a repressed state with the tri-methylation of the lysine-9 residue on histone 3 (H3K9me3). 2. Demethylases from the KDM4 family remove these repressive methyl groups to expose heterochromatin-rich genomic regions. 3. DICER cleaves double stranded RNA (both strands are transcribed) to generate short sequences that then bind to argonaut proteins (e.g. AGO1) to 4. direct the RITS complex back to the original genomic locations where bound H3K9 methyltransferases (EHMT1) re-establish suppression by writing H3K9me3 back to histones. 5. Occupancy of the DNA by chromatin-modifying complexes such as BAF counteract the re-suppression by the RITS complex to allow for transcription of genes in these regions. Genes in red are known to be causative of subtypes of autism, those in yellow are linked to high frequency ASD-SVs in this study and our previous report. PIWIL1 is also a component of RITS analogous to argonaut proteins ^51^, but it is typically expressed in the germline to suppress transposons as well as genes ^52^. Created with BioRender.

Notably, two subtypes of autism are caused by mutations in the proteins that carry out this process. Kleefstra Syndrome is the result of mutations in the H3K9me3 writer *EHMT1* ^34^ and Rett Syndrome is caused by mutations in the *MECP2* gene that codes for a DNA-methylase that modifies the DNA where it contacts H3K9 residues ^35–37^. Autism-like syndromes have also been reported in cases of *de novo* deletions of *AGO1* ^38,39^. We, and others, have identified the H3K9me3 demethylases *KDM4B* and *KDM4C* as risk-genes for ASD (Table S11) ^9^. Here we add to this well-established model the following three transcription factors whose binding sites are enriched for ASD-SVs and participate in the maintenance of heterochromatin.

### SATB1 (Special AT-Rich Sequence Binding Protein 1)

Reports from more than two decades ago established that *SATB1* is associated with H3K9 methylation, heterochromatin, and brain-specific gene regulation ^40^. Later work established that it balances activation and repression states of heterochromatin by regulating acetylation versus methylation of H3K9 ^41^. More recent work determined that it is necessary for *X-ist* mediated X inactivation during embryogenesis in which one X chromosome in each cell of a female is silenced by converting the entire chromosome to a state of constitutive heterochromatin ^42^. Our analysis here indicates that the ASD-SVs are 1.5 times as likely to overlap SATB1 binding sites as expected by chance and is the most common of the three significant binding sites predicted to be disrupted (*p* < 2.7 x 10^-25^, Table S8). In support of our results, haploinsufficiency of *SATB1* itself has been reported to cause an autism-like neurodevelopmental condition, so it stands to reason that disruption of SATB1 target motifs could drive a range of ASD-like conditions ^43^.

### SRSF9 (Serine And Arginine Rich Splicing Factor 9)

In our previous study, we predicted aberrant splicing of the *GRIK2* gene due the presence of an ASD-SV tagging SNP near a known binding site for SRSF9, and confirmed the prediction using RNA-seq data from post-mortem brain tissue from individuals with autism ^5^. In addition to its role as a component of the spliceosome, SRSF9 regulates brain-specific editing of mRNA by the enzyme ADAR2 (also called NUP98) that is necessary for normal neuronal development ^44,45^.

This often occurs at *Alu* retrotransposons, which can affect the stability and splicing of the mRNA transcripts directly, or alter expression by affecting the RITS-mediated formation of heterochromatin by destabilizing siRNAs ^46^ (Figure 2). Our results indicate that individuals with autism should demonstrate disrupted RNA-editing, which is supported by reports of decreased editing in post-mortem brain tissue in those with ASD compared to controls ^47^.

### NUP98-HOXA9 fusion protein

A common non-homologous recombination event between the *NUP98* gene on chromosome 11 (also called *ADAR2*) and *HOXA9* on chromosome 7 can result in a fusion protein that results in the expression of the homeobox locus genes downstream of HOXA9, in addition to many other genes. Although there are no reports of this fusion event in ASD, several lines of evidence support the role of this fusion protein in autism and support our model here. Firstly, as noted above, the NUP98 protein (also called ADAR2) associates with SRSF9 to regulate RITS and neuronal-specific genes. Secondly, the fusion protein along with the protein XPO1 targets heterochromatin ^48^, and thirdly, microdeletions of XPO1 cause ASD-like syndromes ^49^. Finally, one of the ASD-SV tagging SNPs from our analysis that overlaps a NUP98-HOXA9 binding site in ReMap (rs9695393) is adjacent to the microRNA mir-873, known to reduce the expression of ASD risk genes *ARID1B, SHANK3* and *NRXN2* ^50^.

### Summary of Findings and Concluding Remarks

Our results indicate that dysregulation of heterochromatin is at the core of autism spectrum disorder, which directly affects coding-genes that reside in these locations. A Gene Ontology analysis of genes residing in the densest heterochromatin regions of the human genome (gpos100) show significant enrichment for neurodevelopment and immune response whereas those in euchromatin show no significant biological enrichments (Table S9), supporting this hypothesis. These regions are normally repressed by the RITS complex due to the presence of transposons and repetitive DNA elements because they can cause genomic instability. Recent studies have shown that the proteins that regulate heterochromatin are some of the most rapidly evolving in any organism to counter the high mutation rate of heterochromatin itself ^53^. Although RITS likely represents this rapid evolutionary response to counter deleterious mutations, it appears to have also become an efficient means to regulate gene expression and diversity for the genes that reside in these locations. In other words, transposons and repetitive elements in the genome necessitated the evolution of a system to tightly regulate when those genomic regions were allowed to be expressed.

SVs in these regions therefore would be predicted to disrupt this important regulatory process and the evolutionary rate of the genes located within due to a higher mutation rate. Seen in this manner, autism is the phenotypic expression of genetic variants generated within rapidly evolving regions of the genome that are likely directed at the evolution of larger brains in the Hominid lineage as noted previously ^54^. In support of this model, an ASD-SV tagged at rs1957862 in the *LRFN5* gene (Table S2a) is found in nearly ⅓ of individuals in both the Miami and AGPC cohorts we analyzed previously ^5^. This is an ASD susceptibility locus, is involved in synaptic development, and was shown to be differentially marked with H3K9me3 in male individuals with autism when maternally inherited ^55^.

Mechanistically, the observation that heterochromatin may underlie the molecular basis of autism clarifies previous reports of other chromatin modifiers in the disorder ^3,10,56^. Once the suppressed heterochromatin state has been relaxed by removal of the H3K9 trimethylation, these DNA regions are exposed to ATP-dependent chromatin remodeling complexes such as SWI/SNF (also called BAF), ISWI, and NuRD ^25^. Mutations in subunits important for the function of these complexes have been associated with or are causal of autism ^10,11,56^. We also found numerous ASD-SVs in these genes in our previous work and report them here (Table S11). These ATP-dependent chromatin remodelers act to balance suppression and activation of heterochromatin because their depletion results in the rapid return to the quiescent state ^57^. Therefore, any mutations that alter the levels or efficiency of these chromatin-modifying complexes will affect the expression of the genes in those locations as well as the state of heterochromatin in adjacent genomic regions, again emphasizing the importance of heterochromatin in the etiology of ASD.

Our analysis suggests that the primary cause of autism is the dysfunction or dysregulation of the maintenance of heterochromatin. As a result, there is an increase in mutation and generation of structural variation in the genes within the heterochromatin-rich regions of the genome. As mentioned above, dense heterochromatin is enriched for genes involved in brain development and immune response (Table S9) and therefore, a Gene Ontology analysis of the coding genes that harbor ASD-SV and are regulated by SATB1, SRSF9, and NUP98-HOXA9, but do not include chromatin-modifying genes should represent the ultimate cause of different ASD phenotypes (Table S10, Figure 3A). In agreement with this, we found a more than 10-fold significant enrichment for genes involved in *retinal ganglion cell axon guidance* that could explain the observation of reductions in the retinal nerve fiber layer in individuals with autism ^58,59^. This may also underlie differences in pupil light reflexes in those with autism as it is controlled by cells in this lineage ^60–62^. This provides an opportunity to associate these responses with individuals carrying mutations in the genes we identified here (*ALCAM, EFNA5, SLIT2, EPHA7*, and *ROBO2,* Figure 3B, Table S12-S14). Likewise, the genes harboring ASD-SVs and linked to olfactory bulb development may explain differences in responses to odors in those with ASD and provide yet another diagnostic test ^63^.

**Figure 3.**
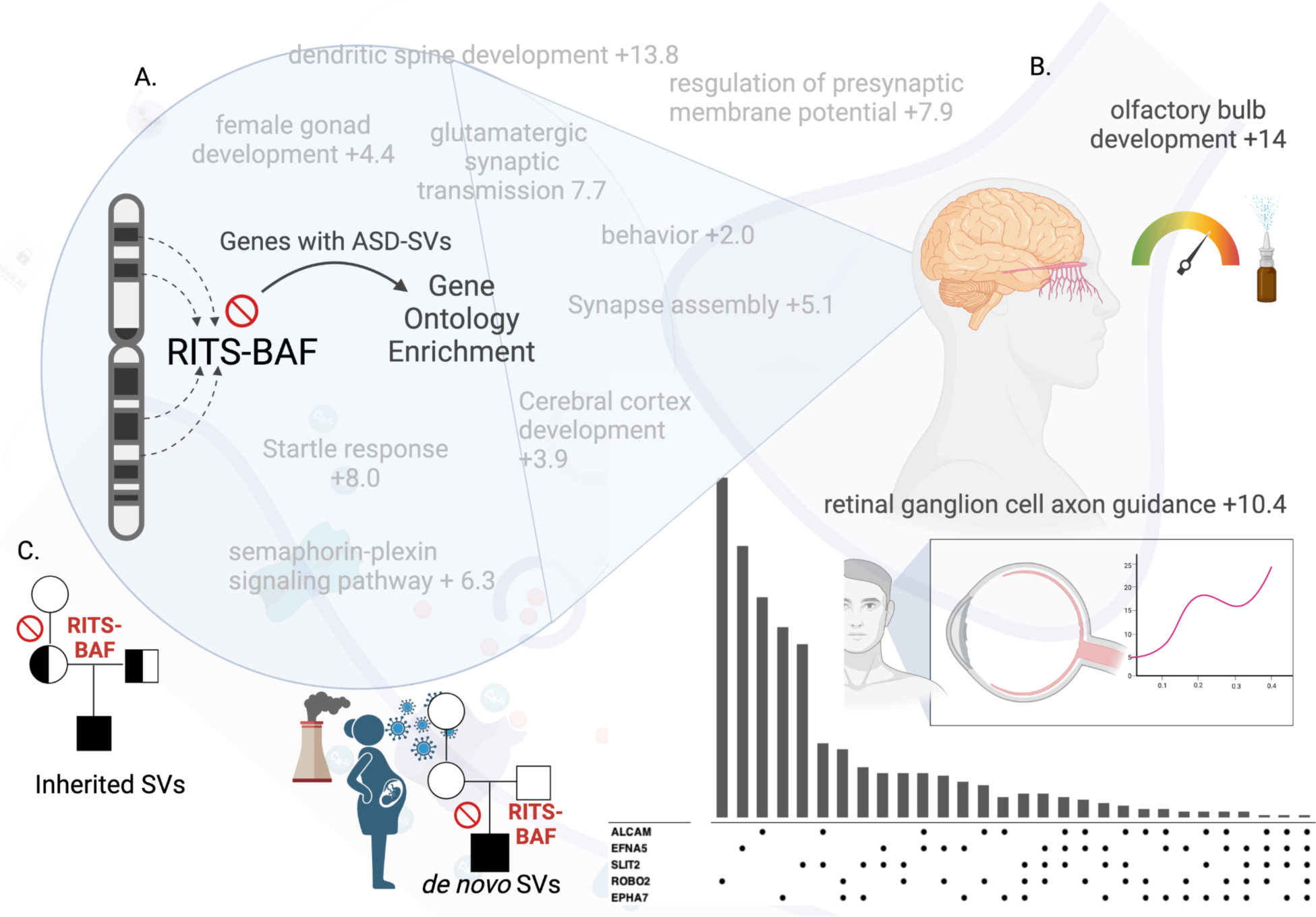
Heterochromatin-centric model for ASD. A) Dysfunctional RITS and/or BAF processes cause structural variants to occur in genes within heterochromatin rich regions. A Gene Ontology (GO) analysis of the genes harboring SVs at high frequency in two cohorts reveal biological processes associated with ASD, such as glutamate signaling and startle response. The individual-level combination of genes affected by SVs generates the heterogeneity of ASD. Numbers indicate fold-enrichment of the ASD-SV genes for those GO biological processes compared to a random set of genes. B) The genes that define these biological processes can be used to develop gene- and individual-focused diagnostic tests. The upset plot (lower right) shows frequency of individuals and the combination of genes with an SV that define the GO category “retinal ganglion axon guidance”. The pupil dilation test for autism may be more precise when accounting for the genetic contributions to the trait. Likewise, the combination of specific genes with SVs in an individual may provide an accurate means to understand differences in olfaction associated with ASD. C) Our results suggest a dysfunctional RITS-BAF system underlies ASD by generating SVs that then segregate like any genomic variation from parents to children. This disrupted RITS-BAF pathway may also explain the role of de novo mutations in autism as they would be an indirect measure of heritability, i.e. they are the result of the inherited mutations in RITS-BAF that then produce new mutations. This could be exacerbated by the exposome during development.

Finally, these findings may explain the role of environmental factors and *de novo* SVs in ASD. Several studies have identified a higher burden of *de novo* mutations in individuals with autism compared to typical developing individuals ^9,11^. Although these findings are interesting, these mutations are not inherited and therefore would not contribute to estimates of heritability. However, if the core dysfunction in autism is the proper maintenance of heterochromatin, then its dysregulation could increase the frequency of *de novo* mutations due to the improper suppression of transposons and increased non-homologous recombination. ^64^. *De novo* mutations therefore are an indirect measure of heritability because they are the result of inherited dysfunctional genome (heterochromatin) maintenance. This would explain previous reports of no differences in transposon insertions in parents, probands, and unaffected siblings ^65^ and support our model that ASD is the result of the epigenetic interaction among the heterochromatin-associated coding genes within each individual that were generated by the faulty RITS-BAF complex. Likewise, an epigenetic-driven mechanism of autism allows for contributions from the environment because stressors such as pollution or maternal infection during pregnancy can activate transposons and disrupt epigenetic processes that regulate heterochromatin ^32^ (Figure 3).

## Data Availability

All data are included in the upload of this manuscript. Original data are online or available from the NIH database of genotypes and phenotypes.

https://www.sciencedirect.com/science/article/pii/S2666247722000677

## Acknowledgements

The authors would like to acknowledge the authors of the original publications that generated the data made available on the NIH database of genotypes and phenotypes (Anney et al. 2010 and Wang et al. 2009). We also thank Stanton Martin for his feedback on the original manuscript draft.

## Funding

We would like to acknowledge funding from the Laboratory Directed Research & Development Program of Oak Ridge National Laboratory, managed by UT-Battelle, LCC for the US Department of Energy.

## Credit Contributions

Michael Garvin (Funding acquisition, Conceptualization, Data Curation, Formal Analysis, Supervision, Investigation, Methodology, Software, Visualization, Validation, Writing - original draft, Writing - review & editing). David Kainer (Conceptualization, Formal Analysis, Investigation, Methodology, Validation, Visualization, Validation, Writing - original draft, Writing - review & editing).

## Competing interests

Authors declare no competing interests.

## Data and materials availability

All original genotype data was obtained from the Database of Genotype and Phenotypes project numbers phs000267.v5.p2 and phs000436.v1.p1. The data used in this analysis are presented and available online with our previous publication ^5^.

## Notes

### Competing Interest Statement

The authors have declared no competing interest.

### Funding Statement

Funding was from the Laboratory Directed Research & Development Program of Oak Ridge National Laboratory, managed by UT-Battelle, LCC for the US Department of Energy.

### Author Declarations

This study used information from previously published research openly available to the public.

